# Racial differences in atrial fibrillation-related stroke: A patient-level comparative analysis of UK Biobank and Korean nationwide data

**DOI:** 10.1101/2023.05.25.23290561

**Authors:** Dong-Seon Kang, Pil-Sung Yang, Daehoon Kim, Eunsun Jang, Hee Tae Yu, Tae-Hoon Kim, Jung-Hoon Sung, Hui-Nam Pak, Moon-Hyoung Lee, Gregory Y.H. Lip, Boyoung Joung

**Affiliations:** Division of Cardiology, Department of Internal Medicine, Yonsei University College of Medicine, Seoul, Republic of Korea; Division of Cardiology, CHA Bundang Medical Center, CHA University, Seongnam, Republic of Korea; Liverpool Centre for Cardiovascular Science at University of Liverpool, Liverpool John Moores University and Liverpool Heart & Chest Hospital, Liverpool, United Kingdom; and Department of Clinical Medicine, Aalborg University, Aalborg, Denmark

**Keywords:** East Asians, atrial fibrillation, stroke

## Abstract

**Background:** This study aimed to evaluate the differences in stroke risk and the impact of atrial fibrillation (AF) among racial groups by conducting a patient-level comparative analysis using two nationwide datasets.

**Methods and Results:** This study utilized data from the Korean National Health Insurance Service-Health Screening and UK Biobank, which included participants who underwent health examinations between 2005 and 2012. The primary outcome was a composite of ischemic and hemorrhagic stroke. A total of 446,986 East Asians in Korea, 3904 East Asians in the UK, and 403,240 Caucasians in the UK were analyzed. East Asians in Korea had a higher comorbidity burden compared to both UK groups (*p*<0.001). During the follow-up period, the incidence of AF showed no significant difference between East Asians in Korea and the UK (Log-rank *p*=0.21), while Caucasians had a higher rate (Log-rank *p*<0.001). Incidence rates of the primary outcome per 1000 person-years were 3.78 (95% CI 3.72–3.85) for East Asians in Korea, 0.92 (95% CI 0.64–1.20) for East Asians in the UK, and 1.13 (95% CI 1.10–1.16) Caucasians in the UK. Although there was no difference between the two UK groups (*p*=0.13), the rate was significantly higher among East Asians in Korea (*p*<0.001). This trend consistently observed regardless of AF status or oral anticoagulant use.

**Conclusions:** Based on this patient-level analysis, East Asians in Korea, unlike East Asians in the UK, were more susceptible to stroke compared to Caucasians. This increased vulnerability was partly attributed to their higher comorbidity burden.

**Key messages:** *What is new?:*

- This large-scale study utilizes patient-level data from approximately one million individuals to evaluate stroke risk differences and the impact of atrial fibrillation (AF).
- While East Asians in the UK demonstrated a similar stroke incidence to Caucasians in the UK, East Asians in Korea, who had the greatest burden of underlying cardiovascular disease such as hypertension, dyslipidemia, and heart failure, showed a stroke incidence rate more than three times that of the two UK race groups.
- The impact of AF on stroke was not significantly difference between the East Asians in Korea and Caucasians in the UK.

*What are the clinical implications?:*

- In contrast to East Asians in Korea, East Asians in the UK demonstrated a stroke risk similar to that of Caucasians, underscoring the imperative need for rigorous management of cardiovascular risk factors and lifestyle modification in order to prevent stroke.
- Future studies should investigate how other risk factors, such as extreme weather, dietary habits, and genetic factors, contribute to the differences in long-term prognosis between East Asians and other racial groups.

## Introduction

Atrial fibrillation (AF) increases the risk of death and morbidity from stroke, congestive heart failure, and poor quality of life, which results in a high socioeconomic burden and increasing healthcare costs.^1, 2^ In the United States, AF affects approximately 1% of the adult population and more than 5% of the population aged 65 years or older, and has been associated with a 5.0-fold increase in stroke risk, and a 2.0-fold increase in mortality.^3^ However, reports on AF-related adverse outcomes have predominantly focused on the majority race, Caucasians. While some healthcare registries and community-based studies have reported the relative incidence of AF in Blacks, Hispanics, and Asians compared to Caucasians, it is crucial to consider that these racial groups, as minorities in their respective countries, participate less in cardiovascular research compared to Caucasians.^4–6^ Similar circumstances were observed in stroke studies as well, where several studies were designed to compare Asians with Caucasians, but lacked information on AF, and patient-level analysis was not feasible, resulting in limitations in overcoming inaccurate or incomplete case ascertainment.^7^ A better understanding of race-specific AF characteristics will not only help with medical planning and resource allocation but also guide interventions to address and mitigate disparities. To improve our understanding of incident AF and AF-related adverse outcomes among East Asians and identify differences in outcomes with those of Caucasians, we analyzed the patient-level data of approximately one million individuals from two national cohorts conducted in Korea and the United Kingdom (UK). In particular, we evaluated and compared the risk of stroke and the racial-specific associations of AF with stroke between East Asians and Caucasians in these cohorts.

## Methods

### Korean National Health Insurance Service-Health Screening Cohort

Data from the Korean National Health Insurance Service-Health Screening (K-NHIS-HealS) were released in 2015, and the details of this cohort have been reported previously.^8, 9^ In Korea, 97.1% of the population mandatorily subscribe to the NHIS, with the remaining 3% categorized as people requiring medical aid. The data of K-NHIS-HealS comprises 558,147 Koreans extracted as a 10% simple random sample from all health screening participants aged 40–80 years. Among them, we selected those who had health check-ups between 2005–2012 and were evaluated for eligibility in K-NHIS-HealS. The participants visited the supervisory institution biannually from 2005 to provide data related to baseline medical information, including sociodemographic and anthropometric measurements, comorbidities, family history, investigations (e.g., radiology reports and blood tests), and lifestyle factors (smoking, alcohol intake, and physical activity). They were followed until the time of death, immigration, or the end of the study (December 31, 2013), whichever came first. Death registration based on death certificates is also managed by the National Population Registry of the Korea National Statistical Office.^10, 11^

### UK Biobank Cohort

The UK Biobank’s comprehensive profiles have also previously been reported.^12^ The UK Biobank is a prospective population-based cohort study in the UK that enrolled 502,422 participants aged 37–73 years recruited from England, Scotland, and Wales between March 2006 and August 2010. Participants were invited to the supervisory institutions to provide baseline medical information similar to that of the K-NHIS-HealS survey of Koreans. The participants were followed up until the time of disqualification from the UK Biobank (death or immigration) or to the end of the study (March 31, 2021 [England and Scotland], February 28, 2018 [Wales]), whichever came first. Information on death (date and main causes) was ascertained from the National Health Service Information Centre/National Health Service Central Register Scotland, in which the death registration was based on death certificates.^13, 14^

This study was approved by the institutional review board of Yonsei University Health System (4-2022-1241). Regarding the K-NHIS-HealS, the informed consent requirement was waived because personal identification information was removed after cohort generation. The UK Biobank study has approval from the North West Multicenter Research Ethics Committee (REC approval 21/NM/0157). This research was conducted using the UK Biobank resource under application 77793. Informed consent was obtained by UK Biobank for all participants.

### Covariates and Outcomes

Information on comorbidities in the K-NHIS-HealS and UK Biobank are provided in Table S1, which have been validated in previous studies.^10, 11, 15, 16^ Self-reported data were used to define sex and ethnicity.^17^ Diagnostic/procedural codes associated with hospital encounters were used to determine the presence of comorbidities at baseline. Based on self-report or prescription information collected from linked general practitioner electronic health record data, the use of medications for the treatment of cardiovascular diseases (i.e., oral anticoagulants [OAC], aspirin, and P_2_Y_12_ inhibitors) was assessed.^16^

The primary outcome was defined as a composite of ischemic and hemorrhagic stroke. The secondary outcomes include individual components of the primary outcome. The definition was based on established *International Classification of Disease, Tenth revision (ICD-10)* codes from the K-NHIS-HealS and UK Biobank claims data (Table S2). Related death records were also utilized to define the outcomes.^16, 18^ These codes were in the primary position, and in addition, the procedure codes for concomitant brain imaging were required. We investigated the risk of initial rather than recurrent events in these participants.^19, 20^

### Selection of Participants

A flow diagram of the participant selection process is shown in Figure 1. In the K-NHIS-HealS and UK Biobank data, 457,510 and 502,422 participants, respectively, aged 18 years or older were identified. This study excluded 8291 (4706 in K-NHIS-HealS and 3585 in UK Biobank) participants who died within 180 days of cohort registration or AF diagnosis. In addition, participants with the following conditions or medical history at baseline were also excluded: 1) AF; 2) mitral valve stenosis or history of valve surgery; 3) missing data for body mass index; 4) missing data for anemia. Hence there are no variables with missing data. To confirm the AF history, we applied a look-back period of three years in both cohorts. After excluding 49,902 participants from the UK Biobank who did not report their race as ‘British White’ or ‘Chinese’, finally, 446,986 East Asians from the K-NHIS-HealS and 3904 East Asians and 403,240 Caucasians from the UK Biobank were included in the analysis.

**Figure 1.**
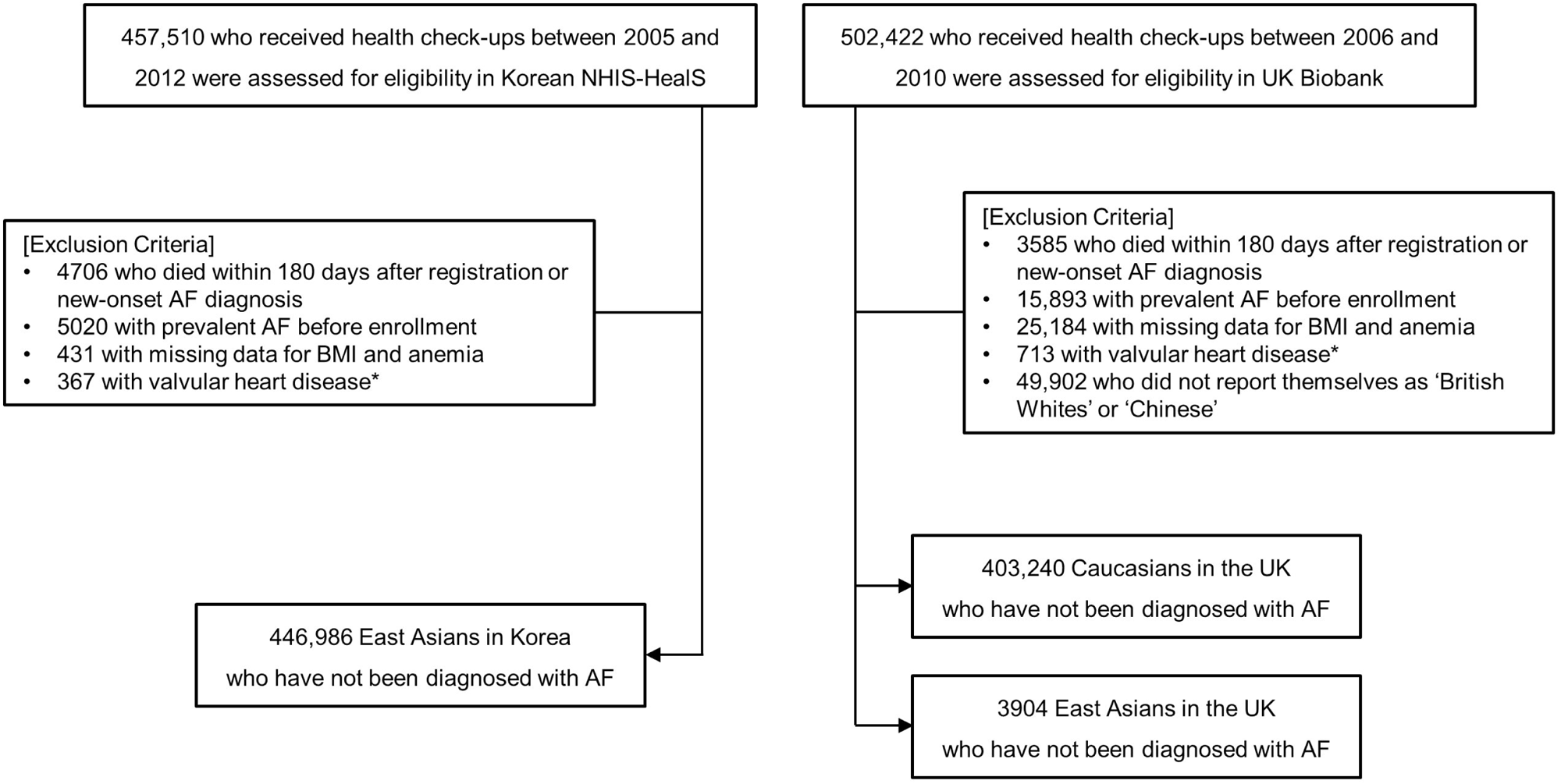
Flow chart of enrollment and analysis of the participants *Defined as a patient with mitral valve stenosis or history of valve surgery AF, atrial fibrillation; BMI, body mass index; K-NHIS-HealS, Korean National Health Insurance Service-Health Screening

### Statistical Analysis

Descriptive statistics were used to depict the baseline characteristics of participants. The incidence rate of the primary and secondary outcomes was calculated as the number of events divided by the total person-years (total of clinical follow-up) and reported per 1000 person-years by race. AF was considered as a time-dependent variable because previous studies had demonstrated that AF duration is associated with the risk of stroke.^19^ Therefore, with respect to participants with AF, they were included in the analysis as those without AF from baseline to the date of AF diagnosis. Cumulative incidence curves were constructed to compare the AF incidence among the three racial groups. To assess the impact of AF on the outcomes, the rate difference was calculated by subtracting the IRs of participants with and without AF, and the hazard ratio (HR) of AF was calculated using multivariable Fine and Gray competing risk regression modeling stratified. In this analysis, East Asians in the UK were excluded due to the insufficient occurrence of events. All-cause death was considered as a competing event. Schoenfeld residuals were used to assess whether the proportional hazards assumption was satisfied and no violations were identified. To explore the race-dependent effect of AF on the primary and secondary outcomes, the model was fitted to the whole study population using an interaction term for AF status and race.

In sensitivity analysis, the analysis was conducted only on OAC-naive participants to account for the impact of OAC on the primary and secondary outcomes. Furthermore, if they were prescribed OAC during follow-up, the observation was censored at the date of OAC initiation. All tests were two-tailed, with *p*<0.05 considered to indicate statistical significance. All analyses were performed using R version 4.2.1 (The R Foundation, www.R-project.org).

## Results

### Baseline Characteristics

After exclusions, there were 446,986 East Asians in Korea, 3904 East Asians in the UK, and 403,240 Caucasians in the UK at baseline, with mean (standard deviation [SD]) ages of 56.0 (9.3), 53.1 (8.2), and 57.2 (8.0), respectively (Table 1). Men comprised 53.9% of East Asians in Korea, 45.4% of East Asians in the UK, and 45.5% of Caucasians in the UK. East Asians in Korea had a higher proportion of participants with a history of hypertension, dyslipidemia, heart failure, stroke, and peripheral artery disease than East Asians and Caucasians in the UK (*p*<0.001). Consistently, East Asians in Korea also demonstrated a higher frequency of baseline cardiovascular medication intake, such as aspirin, P_2_Y_12_ inhibitor, statin, and calcium channel blocker, compared to the other two racial groups (*p*<0.001).

**Table 1.** Baseline characteristics of the study participants

| **Characteristics** | **East Asians**<br>**in Korea**<br>**(N=446,986)** | **East Asians**<br>**in the UK**<br>**(N=3904)** | **Caucasians**<br>**in the UK**<br>**(N=403,240)** | ***p* value** |
| --- | --- | --- | --- | --- |
| Age, y | 56.0 (9.3) | 53.1 (8.2) | 57.2 (8.0) | <0.001 |
| Age≥65 | 88,209 (19.7) | 432 (11.1) | 86,058 (21.3) | <0.001 |
| Men | 241,012 (53.9) | 1774 (45.4) | 183,486 (45.5) | <0.001 |
| Body mass index (kg/m<sup>2</sup>) | 24.0 (2.9) | 25.7 (4.2) | 27.4 (4.7) | <0.001 |
| CHA<sub>2</sub>DS<sub>2</sub>-VASc score* | 1.3 (1.3) | 1.0 (0.9) | 1.1 (0.9) | <0.001 |
| **Comorbidities** |  |  |  |  |
| Hypertension | 141,089 (31.6) | 962 (24.6) | 108,394 (26.9) | <0.001 |
| Diabetes mellitus | 36,755 (8.2) | 370 (9.5) | 18,475 (4.6) | <0.001 |
| Dyslipidemia | 111,011 (24.8) | 628 (16.1) | 56,477 (14.0) | <0.001 |
| Heart failure | 13,573 (3.0) | 69 (1.8) | 8656 (2.1) | <0.001 |
| Previous stroke | 21,081 (4.7) | 12 (0.3) | 3922 (1.0) | <0.001 |
| Previous myocardial infarction | 4675 (1.0) | 86 (2.2) | 8582 (2.1) | <0.001 |
| Previous coronary heart disease | 9819 (2.2) | 104 (2.7) | 10 392 (2.6) | <0.001 |
| Peripheral artery disease | 9302 (2.1) | 2 (0.1) | 1642 (0.4) | <0.001 |
| Hyperthyroidism | 11,266 (2.5) | 61 (1.6) | 3944 (1.0) | <0.001 |
| Hypothyroidism | 11,535 (2.6) | 162 (4.1) | 21,943 (5.4) | <0.001 |
| Previous bleeding history | 7868 (1.8) | 199 (5.1) | 20,477 (5.1) | <0.001 |
| Venous thromboembolism | 1709 (0.4) | 43 (1.1) | 9412 (2.3) | <0.001 |
| Chronic kidney disease | 3386 (0.8) | 26 (0.7) | 3898 (1.0) | <0.001 |
| **Medication at baseline** |  |  |  |  |
| OAC | 740 (0.2) | 7 (0.2) | 1159 (0.3) | <0.001 |
| Aspirin | 85,872 (19.2) | 148 (3.8) | 18,433 (4.6) | <0.001 |
| P<sub>2</sub>Y<sub>12</sub> inhibitor | 11,909 (2.7) | 25 (0.6) | 3560 (0.9) | <0.001 |
| Statin | 51,994 (11.6) | 249 (6.4) | 27,485 (6.8) | <0.001 |
| $\beta$ blocker | 76,121 (17.0) | 111 (2.8) | 21,465 (5.3) | <0.001 |
| Non-DHP CCB | 10,052 (2.2) | 14 (0.4) | 1982 (0.5) | <0.001 |
| DHP CCB | 91,517 (20.5) | 127 (3.3) | 16,132 (4.0) | <0.001 |
| Digoxin | 3408 (0.8) | 1 (0.0) | 149 (0.1) | <0.001 |
| ACEi/ARB | 68,883 (15.4) | 184 (4.7) | 23,314 (5.8) | <0.001 |

Data are presented as means (standard deviations) or No. (%).
\*CHA<sub>2</sub>DS<sub>2</sub>-VASc indicates heart failure, hypertension, age ≥ 75 (doubled), diabetes mellitus, previous stroke or transient ischemic attack (doubled), vascular disease, age 65 to 74, female
6 ACEi, angiotensin converting enzyme inhibitor; AF, atrial fibrillation; ARB, angiotensin receptor 7 blocker; CCB, calcium channel blockers; DHP, dihydropyridine; OAC, oral anticoagulant

### Racial Differences in the Primary and Secondary Outcomes

During the follow-up period, the mean (SD) duration was 7.3 (1.5) years for East Asians in Korea, 11.5 (1.2) years for East Asians in the UK, and 11.7 (1.5) years for Caucasians in the UK. Within this period, the number of AF cases observed were 8534, 86, and 20,744 among East Asians in Korea, East Asians in the UK, and Caucasians in the UK, respectively. The cumulative incidence curves for AF indicate a significantly higher rate among Caucasians in the UK compared to both East Asians in Korea and East Asians in the UK (Log-rank *p*<0.001), while no significant difference was observed between the two East Asian groups (Log-rank *p*=0.21, Figure S1).

However, for the primary and secondary outcomes, results showed a different pattern from the incidence of AF. Regarding the primary outcome, East Asians in Korea, with an incidence rate of 3.78 (95% confidence interval [CI] 3.72–3.85) per 1000 person-years, demonstrated a rate approximately three times higher than that of the two racial groups in the UK (per 1000 person-years, 0.92 [95% CI 0.64–1.20] for East Asians in the UK, 1.13 [95% CI 1.10–1.16] for Caucasians in the UK; *p*<0.001; Figure 2A and Table 2). There was no significant difference between East Asians in the UK and Caucasians in the UK (*p*=0.13).

**Figure 2.**
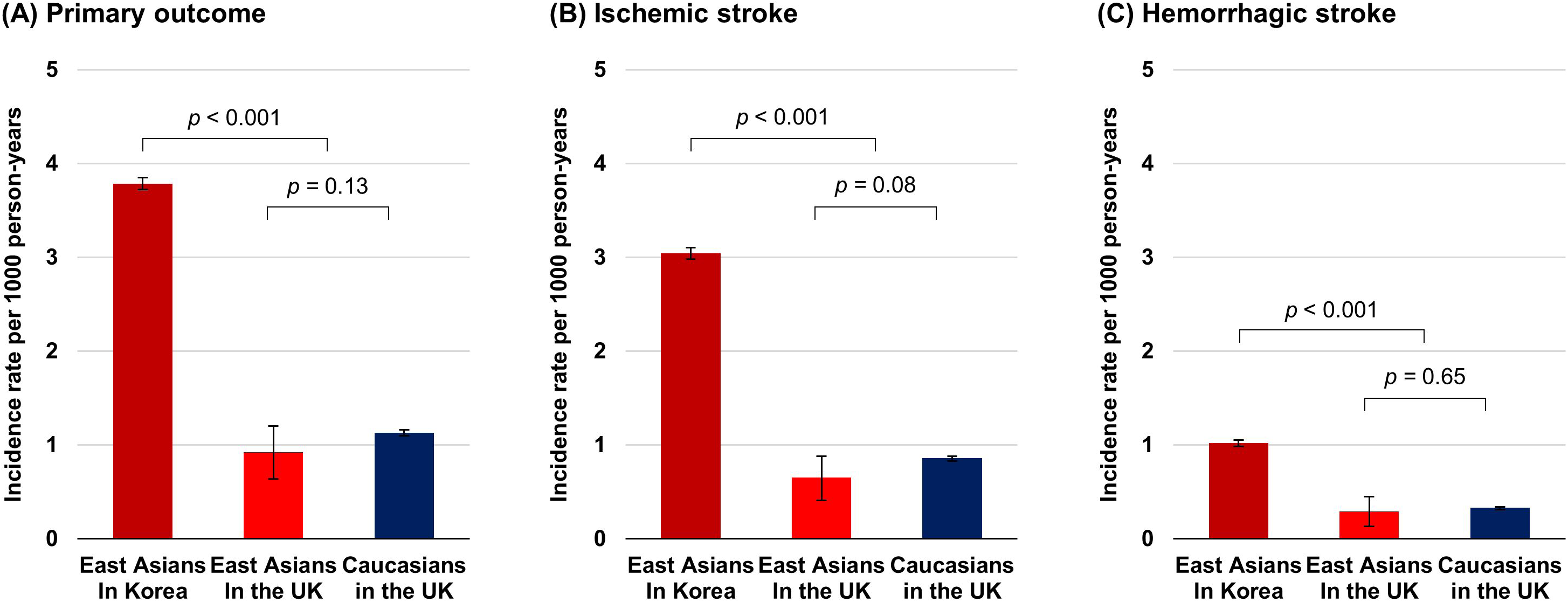
Bar graph of incidence rates for the primary and secondary outcomes in the three racial groups

**Table 2.**
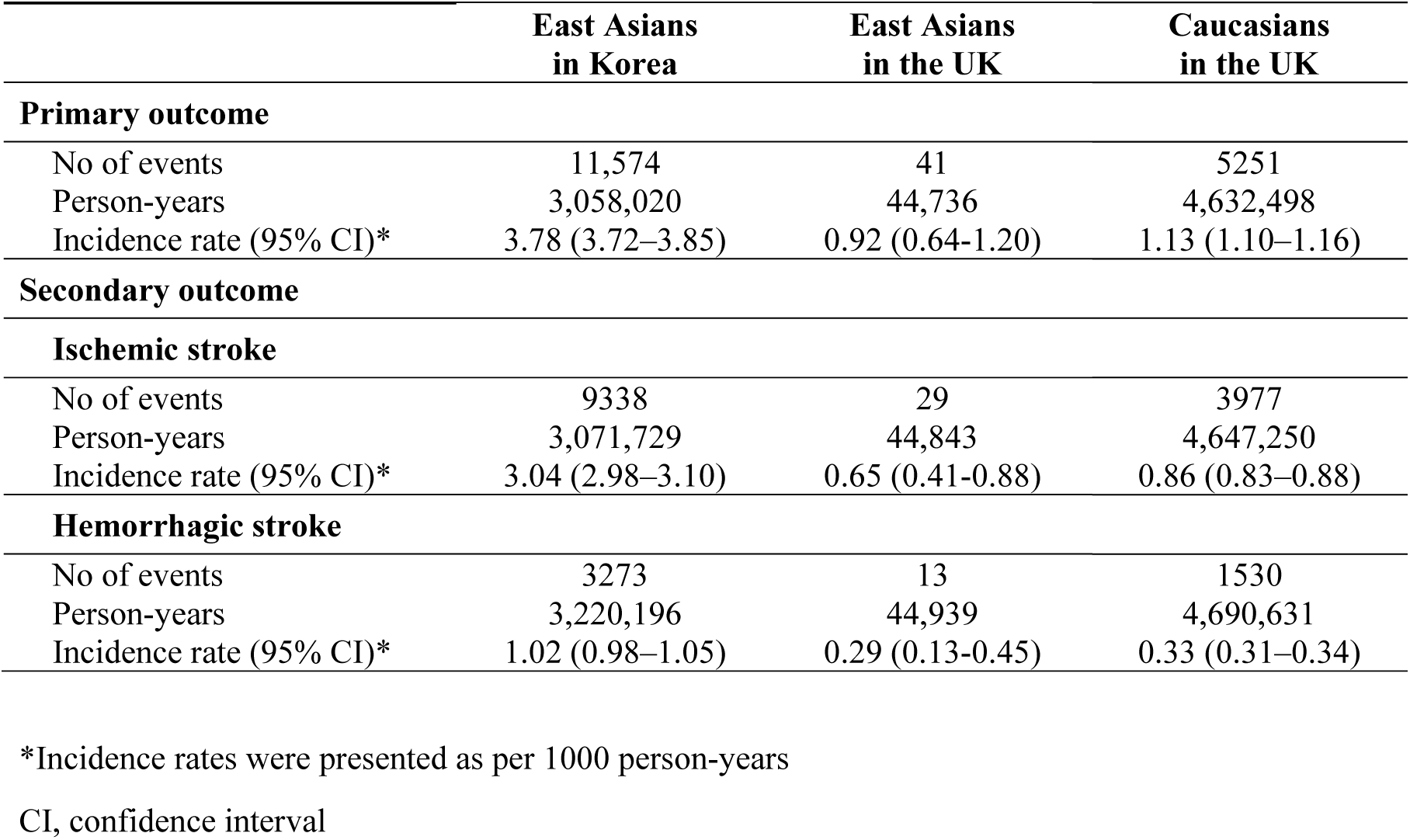
Incidence rates of the primary and secondary outcomes in the three racial groups

Similarly, East Asians in Korea showed an incidence rate of 3.01 (95% CI 2.95–3.07) for ischemic stroke and 1.01 (95% CI 0.98–1.05) for hemorrhagic stroke per 1000 person-years. They exhibited a higher risk when compared to East Asians and Caucasians in the UK for both ischemic (per 1000 person-years, 0.65 [95% CI 0.41–0.88] for East Asians in the UK, 0.85 [95% CI 0.82–0.88] for Caucasians in the UK; *p*<0.001; Figure 2B) and hemorrhagic stroke (per 1000 person-years, 0.29 [95% CI 0.13–0.45] for East Asians in the UK, 0.32 [95% CI 0.31–0.34] for Caucasians in the UK; *p*<0.001; Figure 2C). However, no significant difference was still observed between the two UK groups (*p*=0.08 for ischemic stroke, *p*=0.65 for hemorrhagic stroke).

### Impact of AF on the Primary and Secondary Outcomes

Table S3 summarizes the association of AF with the primary and secondary outcomes during follow-up. East Asians in Korea demonstrated a higher rate of the primary outcome compared to Caucasians in the UK, in both participants with AF (per 1000 person-years, 19.16 [95% CI 17.46– 20.87] for East Asians in Korea, 5.52 [95% CI 5.06–5.98] for Caucasians in the UK; *p*=0.01) and those without AF (per 1000 person-years, 3.66 [95% CI 3.59–3.72] for East Asians in Korea, 1.04 [95% CI 1.01–1.07] for Caucasians in the UK; *p*=0.002). The rate difference for the primary outcome, representing the absolute increase in incidence rate with AF, was 15.51 (95% CI 13.80–17.22) per 1000 person-years for East Asians in Korea and 4.48 (95% CI 4.02–4.94) per 1000 person-years for Caucasians in the UK, respectively. After adjusting for the baseline characteristics presented in Table 1, there was no significant difference in the increased risk of the primary outcome associated with AF between East Asians in Korea and Caucasians in the UK (HR 2.62 [95% CI 2.38–2.87] for East Asians in Korea, 2.85 [95% CI 2.60–3.13] for Caucasians in the UK; *p* for interaction=0.09).

Regarding the secondary outcomes, there was no significant difference in the increased risk of ischemic stroke associated with AF between the two racial groups (HR 2.82 [95% CI 2.55–3.10] for East Asians in Korea, 2.96 [95% CI 2.67–3.28] for Caucasians in the UK; *p* for interaction=0.30). However, the impact of AF on hemorrhagic stroke tended to be lower for East Asians in Korea compared to Caucasians in the UK (HR 1.91 [95% CI 1.58–2.32] for East Asians in Korea, 2.68 [95% CI 2.24–3.20] for Caucasians in the UK; *p* for interaction=0.002).

### Sensitivity Analysis

In the sensitivity analysis, 446,246 East Asians in Korea, 3897 East Asians in the UK, and 402,081 Caucasians in the UK were included as OAC-naive participants. During the follow-up period, 1049 East Asians in Korea, 10 East Asians in the UK, and 2108 Caucasians in the UK started taking OAC. Even if the OAC prescription was regarded as a censoring event, regarding the primary outcome, East Asians in Korea still had a higher rate at 3.76 (95% CI 3.69–3.83) per 1000 person-years, while no significant difference was observed between the two racial groups in the UK (Figure 3 and Table 3). The impact of AF on the primary outcome showed no racial difference (Table S4).

**Figure 3.**
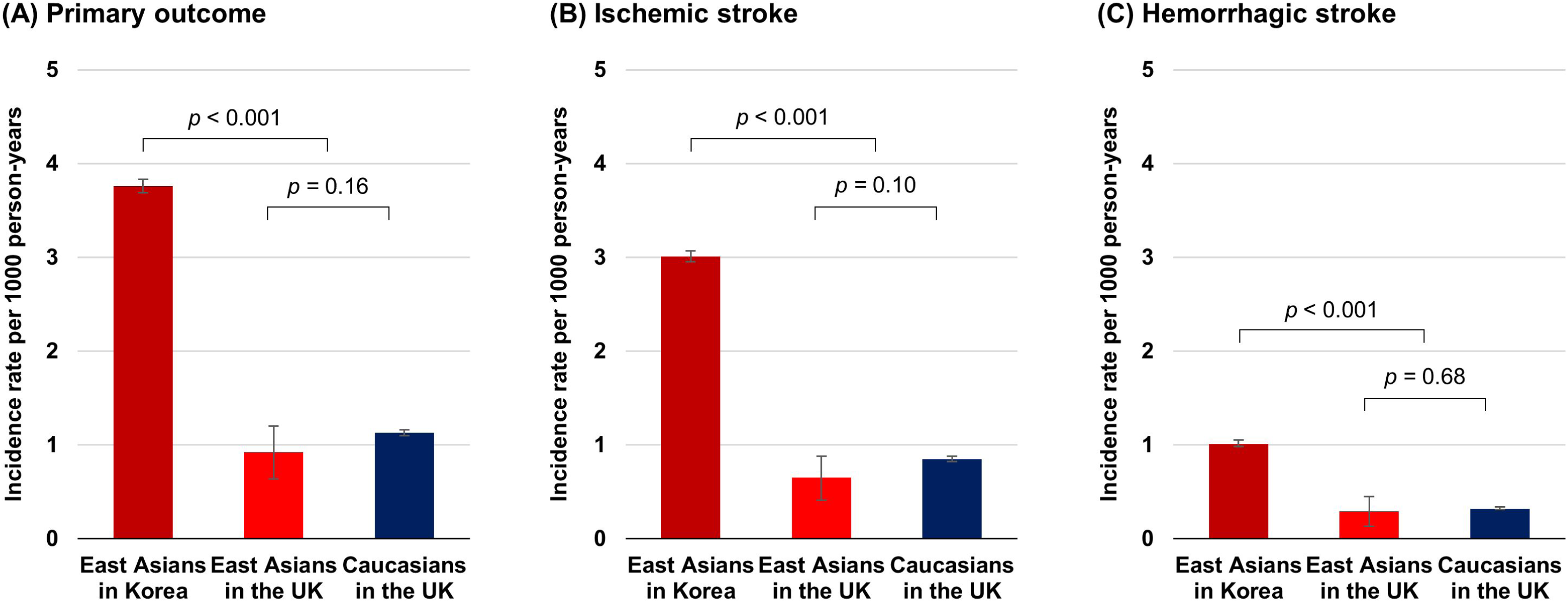
Bar graph of incidence rates for the primary and secondary outcomes among OAC-naive participants

**Table 3.** Incidence rates of the primary and secondary outcomes among OAC-naive participants

|  | **East Asians**<br>**in Korea** | **East Asians**<br>**in the UK** | **Caucasians**<br>**in the UK** |
| --- | --- | --- | --- |
| **Primary outcome** |  |  |  |
| No of events | 11,472 | 41 | 5203 |
| Person-years | 3,053,993 | 44,588 | 4,616,679 |
| Incidence rate (95% CI)* | 3.76 (3.69–3.83) | 0.92 (0.64–1.20) | 1.13 (1.10–1.16) |
| **Secondary outcome** |  |  |  |
| **Ischemic stroke** |  |  |  |
| No of events | 9242 | 29 | 3941 |
| Person-years | 3,067,646 | 44,695 | 4,631,344 |
| Incidence rate (95% CI)* | 3.01 (2.95–3.07) | 0.65 (0.41–0.88) | 0.85 (0.82–0.88) |
| **Hemorrhagic stroke** |  |  |  |
| No of events | 3250 | 13 | 1511 |
| Person-years | 3,213,179 | 44,791 | 4,673,130 |
| Incidence rate (95% CI)* | 1.01 (0.98–1.05) | 0.29 (0.13–0.45) | 0.32 (0.31–0.34) |

\*Incidence rates were presented as per 1000 person-years
CI, confidence interval; OAC, oral anticoagulant

## Discussion

The principal findings of this patient-level comparative analysis of Korean and UK nationwide data that examined the race-specific associations of AF with stroke are as follows: First, East Asians in Korea had the highest burden of cardiovascular comorbidities and drug use. Second, Caucasians in the UK had the highest incidence of AF, with no significant difference observed between East Asians in both countries. Third, the racial group with the highest incidence of stroke was East Asians in Korea, with no significant difference observed between East Asians and Caucasians in the UK. Fourth, the impact of AF on stroke showed no significant difference between East Asians in Korea and Caucasians in the UK.

As far as we are aware, this is the first large-scale patient-level comparative analysis of Asian and Caucasian racial data for AF and AF-related adverse outcomes. Despite a lower incidence of AF compared to Caucasians, the high prevalence of comorbidities, history of drug use, and a stroke incidence three times higher underscore the need for holistic or integrated care, including comorbidity management and lifestyle modifications, particularly in populations residing in East Asia, for the prevention and treatment of AF.^21^ The racial differences in outcomes related to AF are not detailed in current guidelines or expert opinions, partly due to a lack of comparative data.^3, 22^ If racial differences in AF were to be included in these documents by improving East Asian recruitment for clinical trials and registries through active surveillance, this might contribute to the prevention and treatment of AF in East Asians.

Attempts to compare race-related ischemic and hemorrhagic stroke incidences in the general population have been reported.^23^ However, relevant information on specific groups such as East Asians and AF patients are limited due to a lack of methodologically robust studies and limited sample size.^7, 24^ The study, based on substantial individual data from approximately one million people across two national cohorts, and controlled to ensure similar definitions of covariates and outcomes, could reinforce the assertion that populations residing in East Asia are more susceptible to stroke. Some researchers hypothesized differences in the prevalence of cardiovascular risk factors such as hypertension, diabetes mellitus, and obesity to explain Asian-Caucasian differences.^25^ Given its inclusion of East Asians from both Korea and the UK, this study posits that the main contributors to stroke incidence may be environmental and acquired factors such as comorbidities and drug use history, surpassing the influence of race itself. Remarkably, even after adjusting for the effects of comorbidities and drug use history, and accounting for differences in prescription and adherence to OAC,^26^ the impact of AF on stroke was not significantly different between East Asians in Korea and Caucasians in the UK. These consistent findings suggest that the propensity for stroke among East Asians in Korea cannot be explained solely by AF, and also raises questions about other risk factors that may explain the racial differences that were not considered in this study. Racial differences in salt sensitivity could be another plausible explanation, based on experimental observations.^27^ Asians are more likely to have higher salt sensitivity and salt intake than Western populations,^28^ and high salt intake is associated with a significantly increased risk of stroke and total cardiovascular disease,^29^ thus support the vulnerability of East Asians in Korea. Future studies should investigate how other risk factors, such as extreme weather, dietary habits, and genetic factors, contribute to the differences in long-term prognosis between East Asians and other racial groups.

One systematic review has suggested that patients with AF who not receiving anticoagulants were still at overall higher risk of bleeding.^30^ However, few studies have investigated the differences between Caucasians and non-Caucasians, particularly East Asians, in terms of the risk of hemorrhagic stroke following an AF diagnosis.^19, 31, 32^ One study, using data from a prepaid health maintenance organization in the state of California, determined that Blacks, Hispanics, and Asians with AF were at a greater risk of warfarin-related intracranial hemorrhage than Caucasians.^31^ However, only 3.9% of the participants were Asians and the enrollment period between 1995 and 2000 was prior to the introduction of direct oral anticoagulants. Considering that direct oral anticoagulants are generally more effective and safe for East Asians than non-Asians when compared to warfarin, the risk of hemorrhagic stroke in East Asians could be overestimated.^33^ Hence, it is foreseeable that replicating this study’s analysis using contemporary datasets, in an era where direct oral anticoagulants are predominantly used, could prove valuable.

### Limitations

Some limitations exist in our study. First, in this prospective cohort study, there may still be bias due to unidentified confounding factors. For example, the effects of the type and burden of AF that can affect the prognosis of patients, time in therapeutic range among patients treated with warfarin, and period taken to initiate treatment after AF diagnosis were not considered in the study. Second, because the diagnosis was dependent on the ICD-10 codes, a misdiagnosis due to a coding error cannot be excluded. Additionally, as medication use is questionnaire-based information, measurement errors and recall bias may exist. Third, because patients aged 75 years and older were not included in this study, we cannot extrapolate our findings to older population. Fourth, changes in anthropometric and comorbidity variables over time were not accounted for the statistical analysis. Fifth, while K-NHIS-HealS data are considered to be representative of the general population because 10% of all Koreans are randomly selected, the UK Biobank participants are volunteer participants who are relatively healthy compared to the general population, and are likely to be from high socioeconomic areas and women.^34^ Sixth, since all East Asians were recruited from Korea, there is a limit to applying our findings to Asians living in other geographic regions.

## Conclusions

In a patient-level comparative analysis, Caucasians in the UK exhibited a higher incidence of AF than the East Asian groups. Although East Asians in the UK demonstrated a stroke incidence similar to Caucasians in the UK, East Asians in Korea, burdened with higher levels of underlying cardiovascular disease and drug use, showed a stroke incidence rate more than three times that of the two UK race groups. Considering no racial differences in the impact of AF on stroke, these findings suggest that stroke occurrence might be more influenced by environmental and acquired factors than by race itself, thus emphasizing the importance of holistic or integrated management such as comorbidity management and lifestyle modifications, especially among populations residing in East Asia.

## Data Availability

All data and materials of the K-NHIS-HealS and UK Biobank are accessible to the public on their respective homepages (K-NHIS-HealS, http://nhiss.nhis.or.kr; UK Biobank, http://www.ukbiobank.ac.uk). Authorized researchers received anonymous raw data when the independent access subcommittee approved them following the evaluation of all requests for data use by the executive team.

## Acknowledgements

We would like to thank the National Health Insurance Service of Korea for their cooperation. We also would like to thank Dr. Na-hye Kim for her linguistic assistance.

